# Recalibrating SARS-CoV-2 antigen rapid lateral flow test relative sensitivity from validation studies to absolute sensitivity for detecting individuals with live virus

**DOI:** 10.1101/2021.03.19.21253922

**Authors:** Irene Petersen, Alexander Crozier, Iain Buchan, Michael Mina, Jonathan W Bartlett

## Abstract

Testing for SARS-CoV-2 internationally has focused on COVID-19 diagnosis among symptomatic individuals using reverse transcriptase polymerase chain reaction (PCR) tests. Recently, however, SARS-CoV-2 antigen rapid lateral flow tests (LFT) have been rolled out in several countries for testing asymptomatic individuals in public health programmes. Validation studies for LFT have been largely cross-sectional, reporting sensitivity, specificity and predictive values of LFT relative to PCR. However, because PCR detects genetic material left behind for a long period when the individual is no longer infectious, these statistics can under-represent sensitivity of LFT for detecting infectious individuals, especially when sampling asymptomatic populations. LFTs (intended to detect individuals with live virus) validated against PCR (intended to diagnose infection) are not reporting against a gold standard of equivalent measurements. Instead, these validation studies have reported relative performance statistics that need recalibrating to the purpose for which LFT is being used. We present an approach to this recalibration.

We derive a formula for recalibrating relative performance statistics from LFT vs PCR validation studies to give likely absolute sensitivity of LFT for detecting individuals with live virus. We show the differences between widely reported apparent sensitivities of LFT and its absolute sensitivity as a test of presence of live virus. After accounting for within-individual viral kinetics and epidemic dynamics within asymptomatic populations we show that a highly performant test of live virus should show a LFT-to-PCR relative sensitivity of less than 50% in conventional validation studies, which after re-calibration would be an absolute sensitivity of more than 80%.

Further studies are needed to ascertain the absolute sensitivity of LFT as a test of infectiousness in COVID-19 responses. These studies should include sampling for viral cultures and longitudinal series of LFT and PCR, ideally in cohorts sampled from both contacts of cases and the general population.

## Introduction

Testing for SARS-CoV-2 in most countries has, until recently, focused on COVID-19 diagnosis among symptomatic individuals using quantitative reverse transcriptase polymerase chain reaction (PCR) tests. Recently, however, SARS-CoV-2 antigen rapid lateral flow tests (LFT) have been rolled out in several countries for testing asymptomatic individuals in public health programmes and for providing a more rapid, low-cost alternative to PCR in specific contexts.(1)

In contrast to PCR, LFT are primarily being used to identify infectious individuals who do not display classical COVID-19 symptoms, or at least do not use symptomatic testing centres. It is well recognised that pre-symptomatic transmission is a key driver of spread, with many individuals being infectious without displaying classic symptoms - around a third of transmission may be accounted for by this group.(2,3) Validation of LFT for asymptomatic testing has employed several cross-sectional studies and reported conventional diagnostic test accuracy statistics. However, given PCR is testing for any sign of SARS-CoV-2 RNA, which most of the time will be material left over after the individual has stopped being infectious, and that LFT is testing for shedding of viral antigen with the intention of identifying infectious cases, these two tests are not comparable.(4) Therefore, LFT validated against PCR is not reporting diagnostic accuracy statistics against a gold standard of equivalent measurements. Instead, these validation studies have reported relative performance statistics that need recalibrating to the purpose for which LFT is being used.

In most of the validation studies, individuals were tested simultaneously with LFT and PCR, with PCR used as a gold standard i.e., a marker of SARS-CoV-2 infectiousness, when it is a surrogate marker for SARS-CoV-2 infection. Sensitivity was thus evaluated as the ability of LFT to identify the same positive cases as the PCR and the specificity as the ability to identify the same negative cases as the PCR.(5–8) In these studies, it often appears that the LFTs have a low sensitivity, but high specificity. For example, in a pilot study of asymptomatic testing in Liverpool the sensitivity and specificity of the LFT was estimated to be 40.0% (28.5% to 52.4%; 28/70) and 99.9% (99.8% to 99.99%; 5,431/5,434), respectively.(5) In a Danish study, the estimates were 69.7% and 99.5%, respectively.(6) In a Spanish study of close contacts the estimate of sensitivity among asymptomatic individuals was 60% and 80% among symptomatic individuals.(7) An evaluation of a rapid antigen test done by the Centers for Disease Controls and Prevention (CDC) found the sensitivity was 41% (18.4% to 67.1%) and 80% (64.4% to 90.9%) among asymptomatic and symptomatic individuals, respectively (9) and another evaluation by CDC found the sensitivity, relative to PCR, was 36% and 64% amongst asymptomatic and symptomatic populations, respectively. However, when measured against specimens with a positive viral culture, sensitivity rose to 79% to 93% in asymptomatic and symptomatic populations, respectively.(10) These diverging figures have raised some debate about the sensitivity of the LFTs and concerns have been raised about their utility in the context of testing asymptomatic individuals.(11–13)

In order to assist policy makers, we have further investigated the reported sensitivities of LFTs as tests of presence of live virus in the body, and we derive a formula for recalibrating these LFT-to-PCR *relative* performance statistics into *absolute* sensitivity of LFT for detecting individuals with live virus in their body.

### The difference between PCR and LFTs

Before evaluating the results of the validation studies, it is important to understand the biology of SARS-CoV-2 and recognise that the two tests reflect different properties of the infection and have different testing utilities – clinical versus public health. The PCR test is an established laboratory technique which can be used to identify the presence of SARS-CoV-2 by cycles of amplification of viral RNA present in a sample. At each cycle, the RNA doubles and so the test is exquisitely sensitive, potentially able to detect a single RNA fragment in a sample. The more cycles it takes to detect an infection the less RNA there is in a sample, and the repetition of cycles is usually abandoned at 35. The point at which RNA is detected or cycling abandoned is called the cycle threshold (Ct).(14) So, lower Ct values reflect higher levels of virus RNA. Due to the amplification step, PCR can detect RNA in quantities much lower than the limits of detection by virus culture, which is typically recognised as the proxy for infectiousness.(15) As such, while PCR is highly sensitive it is poorly specific for *infectiousness*, the fundamental characteristic of communicable disease and a critical consideration in the control of SARS-CoV-2 transmission. It is important to note that PCR is still the gold-standard for diagnostic testing - if the test is conducted shortly after an individual develop symptoms, it is highly likely a positive PCR result equates to an infectious case as most people who develop symptoms are infectious just before and for a median 5 days after symptom onset.(16) A large systematic review on the duration of viral shedding and infectiousness found mean duration of SARS-CoV-2 RNA shedding in upper respiratory tract is 17 days (95% CI 15·5–18·6). However, there is large variation and many studies did not take into account that shedding may have started several days before individuals received their first positive test results. Therefore, in many people RNA shedding may occur for three weeks or more. Most people infected with SARS-CoV-2 are infectious for 4–8 days and no studies have detected live virus beyond day 9 of illness. (17) Because of the prolonged RNA shedding and relatively short infectious time window, we will expect only to find 50% or less in an asymptomatic sample to be within the infectious time window when they test positive on PCR. This discrepancy is also recognised by Public Health England guidance recommending individuals not to be tested with PCR tests within 90 days of exiting isolation because they may remain positive for a long time.(18)

LFTs are an established technology, for example used in pregnancy test kits, that was repurposed for detecting SARS-CoV-2 surface proteins (antigens) that are present in the samples from infectious people. The tests do not require laboratory processing and results are provided within 10-30 minutes. (19) LFTs have a high specificity and are unlikely to provide positive results long after the infectious period, in contrast to PCR.(1,20)

When performing cross-sectional testing in a population, the ratio of currently infectious to post-infectious cases is expected to change with the phase of the epidemic curve.(21) This ratio is higher when the epidemic is surging and lower when it is shrinking or in steady state because the time course of infectiousness within infected individuals is asymmetrical (front-loaded). Where there is sustained community transmission there will be a significant proportion of individuals who are beyond the infectious period, but still shedding RNA which will be detected by PCR, but unlikely to be detected by the LFTs.(21)

### Calibrated sensitivity of LFTs

Based on the information about the biology of SARS-CoV-2, the dynamic of the epidemic and data on the performance of the PCR and LFTs, we illustrate how the reported sensitivity estimates in cross-sectional validation studies likely underestimate the sensitivity of the LFTs in terms of detecting individuals who are carrying live virus material at the time of the test.

We derive a formula for the ***apparent relative sensitivity*** (1) of the LFT from a cross-sectional validation study when using the PCR as the reference test, which we then re-arrange for calculating a ***calibrated absolute sensitivity*** (2) of the LFT for detecting individuals with live virus.

Let D denote an individual’s infection status, with D=0 for never infected or previously infected but no longer RNA shedding, D=1 for infected and with live virus and D=2 for post-infectious (still RNA shedding, but no longer with live virus in their body). Let T_PCR_ and T_LFT_ denote the PCR and LFT outcomes for an individual, with + and – for positive and negative results. We assume the two test results depend only on the individual’s true infection status D. To simplify this further we assume the PCR test never tests positive in those who have never been infected (D=0) and PCR always tests positive in those with D=1 or D=2. We additionally assume that LFT never tests positive in post-infectious (D=2) individuals. Under these assumptions we can derive the following formula for the *apparent relative sensitivity* of the LFT from a cross-sectional validation study when using the PCR as the reference test:

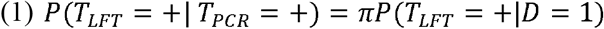

where 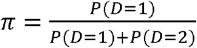 denotes the proportion of individuals with live virus in their body among those who are RNA shedding (D=1 or D=2). This shows the sensitivity relative to PCR will appear markedly lower than its sensitivity for detecting those who currently carry live virus in their body (D=1). Re-arranging the formula shows the calibrated absolute sensitivity of LFTs for detecting those individuals with live virus is:

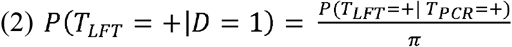

### Limitations

We note that PCR is likely to identify potential infectious individuals slightly earlier than LFTs, but due to the exponential growth of the infection this time window is quite short (perhaps less than 24 hours) and therefore is not included in our calibration. We recommend further longitudinal series of LFT and PCR to determine the time difference between turning positive on PCR and LFT, and how this relates to symptom onset and transmission.

Likewise, we acknowledge that the sensitivity of the tests may be affected by sampling error and experience of the person performing the sampling and the test. These uncertainties are not taken into account in our calibrations, but they are discussed in the reports from Liverpool and by Peto and colleagues.(5,20)

### Calibrated absolute sensitivity for detecting individuals with live virus in their body

Table 1 illustrates the relationship between the proportion of individuals with live virus among those who are shedding RNA *(π*), the apparent sensitivity and calibrated sensitivity of LFTs.

**Table 1.**
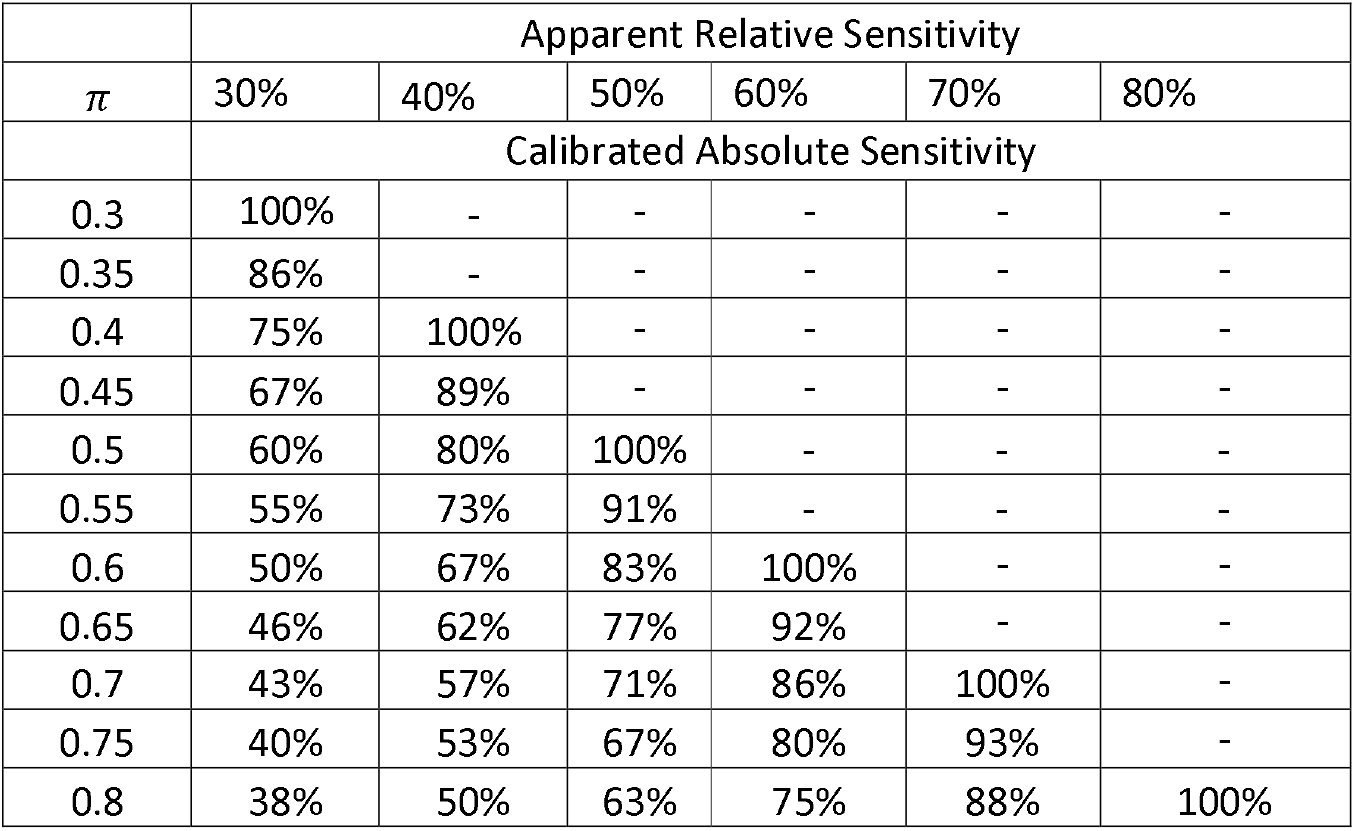
Calibrated sensitivity (%) of LFT to detect individuals with live virus in their body by different values of *π* and apparent sensitivity estimates. - indicates scenario not possible under the given assumptions.

From Table 1 it can be seen that the upper limit (100%) of the calibrated absolute sensitivity is reached when *π* and apparent relative sensitivity are equivalent. For example, in a sample where under half of the individuals still have live virus in their body a validation study with PCR test as the reference test can never reach an apparent sensitivity of the LFTs of more than 50%. On the other hand, in a study which includes symptomatic individuals the proportion of individuals with live virus in their body in the sample *π* is likely to be much higher and thus we would expect the apparent sensitivity to be higher. This is what was observed in the validation studies in Denmark, Spain and US.(6,7,9) As mentioned above the proportion of individuals with live virus in a sample also varies over time and across locations. Thus, we may find the apparent validity of the *same* type of LFTs also varies substantially between studies carried out in different location and at different times. Hence, a large variation of the apparent sensitivity has been observed in empirical studies carried out so far.(5–7,9)

One outstanding question remains; what is the actual sensitivity of the LFTs in terms of identifying individuals with live virus in their body? With knowledge of the biology of the virus and information about the local developments of the pandemic we can calculate calibrated sensitivities of the test. For example, using the data from the Liverpool validation study (5) we would expect *π* to be smaller than 0.5 based on our knowledge of the virus. It may have been even smaller at the time of the study as the epidemic was shrinking in Liverpool by December 2020. This suggests that the apparent relative sensitivity of 40% found in Liverpool may result in a calibrated absolute sensitivity above 80%. As we will never know the exact value of *π* in these validation studies we cannot provide an exact value for the calibrated absolute sensitivity.

### Further validation of LFTs

As LFTs are becoming more widely used in schools and work places, it is important that health professionals and the public have clear information about the operating characteristics of the tests. We have demonstrated that the calibrated absolute sensitivity to detect live virus is likely high with LFT. To improve our understanding of their characteristics, longitudinal studies where individuals, ideally contacts of cases, are tested daily by LFT and PCR would help to further to and the time differences in terms of turning positive between PCR and LFT. Further validation of the LFTs could be performed against live viral cultures as it was done by Prince-Guerra (10) and Pickering et al.(22), or, ideally, against a gold-standard antigen test with proven accuracy for detection of not only live virus, but also infectious individuals. Some validation studies have sought to use specific Ct cut-offs when comparing the performance of PCR and LFTs (20), but differently calibrated PCR systems mean that Ct levels can’t easily be compared between studies, and the values do not always indicate the same level of virus in a sample between laboratories.(4,13)

Misleading criticisms of LFT for apparent low sensitivity have failed to take the viral biology and epidemiology into account and we believe have reached the wrong conclusions.(11,12,23) This has confused policy making and damaged public trust in LFTs, despite the need for better tools to control transmission of SARS-CoV-2. (4) It is our hope recalibrated absolute sensitivity statistics will assist policy making and help to build public confidence in LFT as a tool to aid COVID-19 resilience and recovery.

## Conclusion

In this study we investigated validation studies of LFT and showed the pitfalls of reporting sensitivity values relative to PCR as if they were absolute values measured against a gold standard test. In most samples of asymptomatic individuals, we would expect the less than half of PCR positive individuals to be carrying live virus. A well-performing test of presence of live virus would therefore have an apparent relative (to PCR) sensitivity never exceeding 50%. Recalibrating and an apparent relative sensitivity of 50%, on average we would expect an approximate absolute sensitivity of over 80% in testing for individuals with live virus. Future studies might improve this calibration further using series of daily repeated PCR and LFT among substantial cohorts drawn from the general population, and viral cultures from LFT samples.

## Data Availability

The figures used in this study is based on published material.

## Formula derivation

To derive the calibrated sensitivity formula, we have:

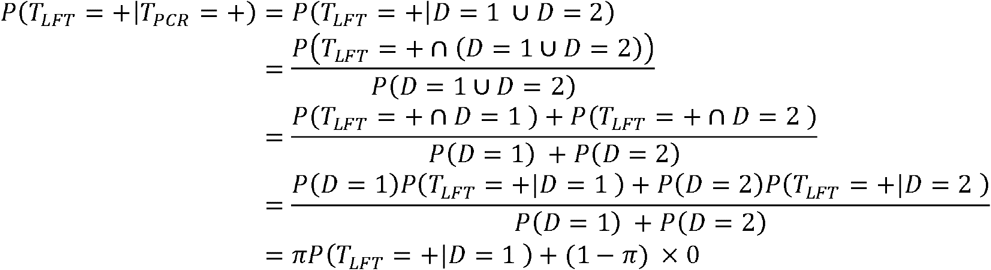

where in the first line we use our assumption that PCR never tests positive in D=0 individuals and always tests positive in D=1 or D=2 individuals, such that testing positive by PCR is equivalent to the event D=1 or D=2 and we use the fact that D=1 and D=2 are mutually exclusive.

